# Early pregnancy metabolomics and risk of offspring heart defects: a matched case-control study

**DOI:** 10.64898/2026.05.08.26352715

**Authors:** Katerina Nastou, Filip Ottosson, Amalie Bøggild Schmidt, Giulia Corn, Frank Geller, Sanne Grundvad Boelt, Nadia MacSween, Jan Wohlfahrt, Marie Lund, Mads Melbye, Madeleine Ernst, Bjarke Feenstra

**Author notes:** equal contribution (Katerina Nastou, Filip Ottosson).

## Abstract

Congenital heart defects (CHDs) are the most common congenital malformations and often arise from perturbations during early embryonic development. Maternal metabolic disturbances in early pregnancy may contribute to CHD risk, but evidence from early first-trimester metabolomics studies is limited. We conducted an untargeted metabolomics case-control study using early first-trimester maternal plasma samples (gestational weeks 4–10) from the Danish National Birth Cohort. Metabolite profiling was performed via liquid chromatography-tandem mass spectrometry (LC-MS/MS) on 160 matched CHD case-control pairs (320 total samples). Conditional logistic regression and interaction analysis were used to identify metabolites associated with CHD risk or specific cardiac phenotypes. A total of 1,471 metabolite features were measured with 69 metabolites being associated with CHD at nominal significance (p < 0.05). These included a desaturated analog of sphingosine-1-phosphate (S1P), isoleucylproline and an arginine related metabolite. However, after false discovery rate correction for multiple testing no metabolites remained significant. While these findings do not preclude that subtle metabolic variation may exist in early pregnancy among CHD cases, they also underscore the challenges of biomarker discovery in this context. This work highlights the potential of early-pregnancy metabolomics for CHD biomarker discovery, and points toward more targeted future studies with improved sample collection protocols, pre-specified pathway panels, and phenotype-homogeneous analyses to better capture the subtle metabolic variation that may underlie CHD risk.

## Introduction

Congenital heart defects (CHDs) are structural abnormalities of the heart or intrathoracic vessels occurring during cardiogenesis before the 10^th^ gestational week^1^ and represent the most common congenital anomaly (∼1% of births)^2^ accounting for ∼30% of all anomalies^3^. In Denmark, the prevalence is similarly around 1%, corresponding to approximately 500-600 affected infants annually^4^. Beyond the immediate impacts on neonatal health, CHDs can lead to lifelong health challenges and healthcare needs^5^, underscoring their public health importance.

CHD etiology is multifactorial, involving genetic and environmental factors^6^. Among maternal exposures, pre-gestational diabetes is one of the strongest established maternal modifiable risk factors, associated with an estimated 4 times increased relative risk of CHD in the fetus^7^, a risk elevated across CHD phenotypes. Other factors, including teratogenic medications^8,9^, maternal obesity and nutritional factors, such as folate status, have also been linked to CHD risk. However, for folate status evidence for specific subtypes remains incomplete^10^. Other factors such as smoking, alcohol, advanced age, and different pregnancy complications have also been explored as potential contributors to CHD risk^11–13^.

Metabolomics offers an opportunity to investigate biochemical pathways that may connect maternal exposures and physiology to fetal cardiac development. Prior maternal metabolomics studies of CHDs have identified alterations in lipid, amino acid, and glucose metabolism in mothers carrying fetuses with a CHD^14–19^, but findings have been heterogeneous and no individual metabolite or panel of metabolites has been reproducibly validated as a predictor of CHDs across studies^20^. Key sources of heterogeneity include variations in sample timing, biospecimen type, analytical platforms and CHD phenotyping. Thus, while maternal metabolomic profiling is a compelling tool, further research with well-designed studies is needed to clarify its utility in CHD risk assessment.

Timing of maternal sample collection is critical, as the first trimester of pregnancy represents the key developmental window for the fetal heart. Cardiac morphogenesis begins in the 3^rd^ gestational week and is largely completed by about the 10^th^ gestational week, by which time the basic heart chambers and connections have formed^21^. Therefore, metabolic disturbances in the mother during this early period are biologically relevant to CHD origins and may be informative for early risk stratification.

Modifiable risk factors account for approximately one third of CHDs^22^, underlining a significant potential for prevention. Given the contradictory evidence and current knowledge gaps, we conducted an untargeted metabolomics matched case-control study in early first-trimester maternal plasma samples within a large prospective cohort. We aimed to determine whether maternal metabolic profiles in early pregnancy differ between pregnancies that result in offspring with a CHD and those that result in offspring without a CHD. Specifically, our study employed a liquid chromatography-mass spectrometry (LC-MS/MS) platform to profile metabolites in plasma samples from mothers in the Danish National Birth Cohort (DNBC), comparing 160 CHD case pregnancies to 160 matched control pregnancies. We hypothesized that differences in maternal metabolites could be detected between case and control pregnancies that would allow us to pinpoint specific modifiable risk factors to potentially prevent CHDs.

## Results

### Sample Selection and Quality Control

A total of 210 matched case-control pairs (420 maternal plasma samples) were initially selected from the Danish National Birth Cohort for metabolomics analysis. After quality control (QC), 367 samples remained. To preserve the matched study design, samples lacking a corresponding case or control after QC were excluded, resulting in a final dataset of 160 matched pairs (320 samples) for downstream analysis.

### Clinical Characteristics

Clinical characteristics of the 320 mothers and pregnancies included in this study are summarized in *Table 1*. As expected, due to the matching, the distributions of maternal age and gestational age at sampling were nearly identical between cases and controls. Other variables, such as maternal BMI, smoking, and alcohol consumption prior to pregnancy onset, were comparable between groups and included in covariate-adjusted models. The only statistically significant difference between the two groups is on birth weight. The lower birth weight among cases was expected, as CHD has consistently been associated with reduced fetal growth and lower birth weight at delivery^23^.

**Table 1.** Characteristics of the study sample in cases and controls. Continuous variables (maternal age, BMI, birth weight) are presented as mean (SD) and compared between cases and controls using Welch’s t-tests; categorical variables are shown as n (%) and compared with chi-square tests (smoking, offspring sex, parity) or Fisher’s exact tests with simulated p-values for sparse multi-level variables (region, alcohol). All tests are two-sided. *Region: Region of residence at pregnancy onset; Parity: Number of children from the same women: 1=first child, 2=second child, 3=third child, 4=fourth child, 5=fifth child; BMI: BMI at pregnancy onset; Smoking: Smoking prior to pregnancy onset; Alcohol: Alcohol use prior to pregnancy onset measured in alcohol units per week*.

| Variable | Level | Controls (n = 160) | Cases (n = 160) | p-value |
| --- | --- | --- | --- | --- |
| Region, n (%) | Hovedstaden | 41 (25.6%) | 30 (18.8%) | 0.612 |
|  | Midtjylland | 42 (26.2%) | 41 (25.6%) |  |
|  | Nordjylland | 24 (15.0%) | 26 (16.2%) |  |
|  | Sjælland | 21 (13.1%) | 24 (15.0%) |  |
|  | Syddanmark | 32 (20.0%) | 39 (24.4%) |  |
| **BMI, mean (SD)** | – | 24.64 (9.40) | 25.31 (9.78) | 0.533 |
| **Smoking, n (%)** | No | 118 (73.8%) | 121 (75.6%) | 0.434 |
|  | Occasionally | 21 (13.1%) | 18 (11.2%) |  |
|  | Daily <15 | 15 (9.4%) | 19 (11.9%) |  |
|  | Daily ≥15 | 6 (3.8%) | <5 (<3%) |  |
| **Alcohol intake, n (%)** | 0 | 96 (60.0%) | 94 (58.8%) | 0.911 |
|  | >0 & ≤ 1 | 45 (28.1%) | 50 (31.3%) |  |
|  | >1 & ≤ 2 | 14 (8.7%) | 12 (7.5%) |  |
|  | >2 | 5 (3.1%) | <5 (<3%) |  |
| **Offspring sex, n (%)** | Female | 76 (47.5%) | 73 (45.6%) | 0.823 |
|  | Male | 84 (52.5%) | 87 (54.4%) |  |
| **Parity, n (%)** | 1 | 63 (39.4%) | 72 (45.0%) | 0.688 |
|  | 2 | 67 (41.9%) | 61 (38.1%) |  |
|  | 3 | 27 (16.9%) | 24 (15.0%) |  |
|  | 4 | <5 (<3%) | <5 (<3%) |  |
|  | 5 | <5 (<3%) | <5 (<3%) |  |
| **Gestational week at sampling, n (%)** | 4 | <5 (<3%) | <5 (<3%) |  |
|  | 5 | 5 (3.1%) | 5 (3.1%) |  |
|  | 6 | 23 (14%) | 23 (14%) |  |
|  | 7 | 37 (23%) | 37 (23%) |  |
|  | 8 | 40 (25%) | 40 (25%) |  |
|  | 9 | 31 (19%) | 31 (19%) |  |
|  | 10 | 23 (14%) | 23 (14%) |  |
| **Maternal age, mean (SD)** | (years) | 29.4 (3.9) | 29.5 (4.0) | 0.937 |
| **Birth weight, mean (SD)** | (grams) | 3,666 (588) | 3,436 (705) | 0.002 |

### Associations of metabolites with CHD

We assessed associations between maternal metabolite levels and CHD status using conditional logistic regression (CLR), stratified by matched pairs, and adjusted for covariates. Each metabolite was analyzed in a separate univariate model that included the following covariates: sex, maternal pre-pregnancy BMI, gestational age at blood draw, maternal smoking prior to pregnancy onset, alcohol use prior to pregnancy onset, and parity.

A total of 1,471 metabolites were tested. Of these, 69 metabolites showed nominal associations with CHD status (p < 0.05); however, none remained significant after applying false discovery rate (FDR) correction. Among the 69 nominally significant metabolites, 10 contained MS2 (tandem mass) fragmentation spectra, and 9 of these could be annotated to the chemical class level (*Supplementary Table 1*). The annotated compounds included one sphingolipid (a desaturated analog of S1P), four amino acids or derivatives (including a possible structural analogue of arginine, *Supplementary Figure 1*), one dipeptide (annotated as isoleucylproline), two organoheterocyclic compounds and one compound annotated as organic acid or derivative thereof. More detailed information about the chemical structural information retrieved for each compound can be found in Figure 1, Supplementary Table 1 and *Supplementary Figures 1–3*.

**Figure 1.**
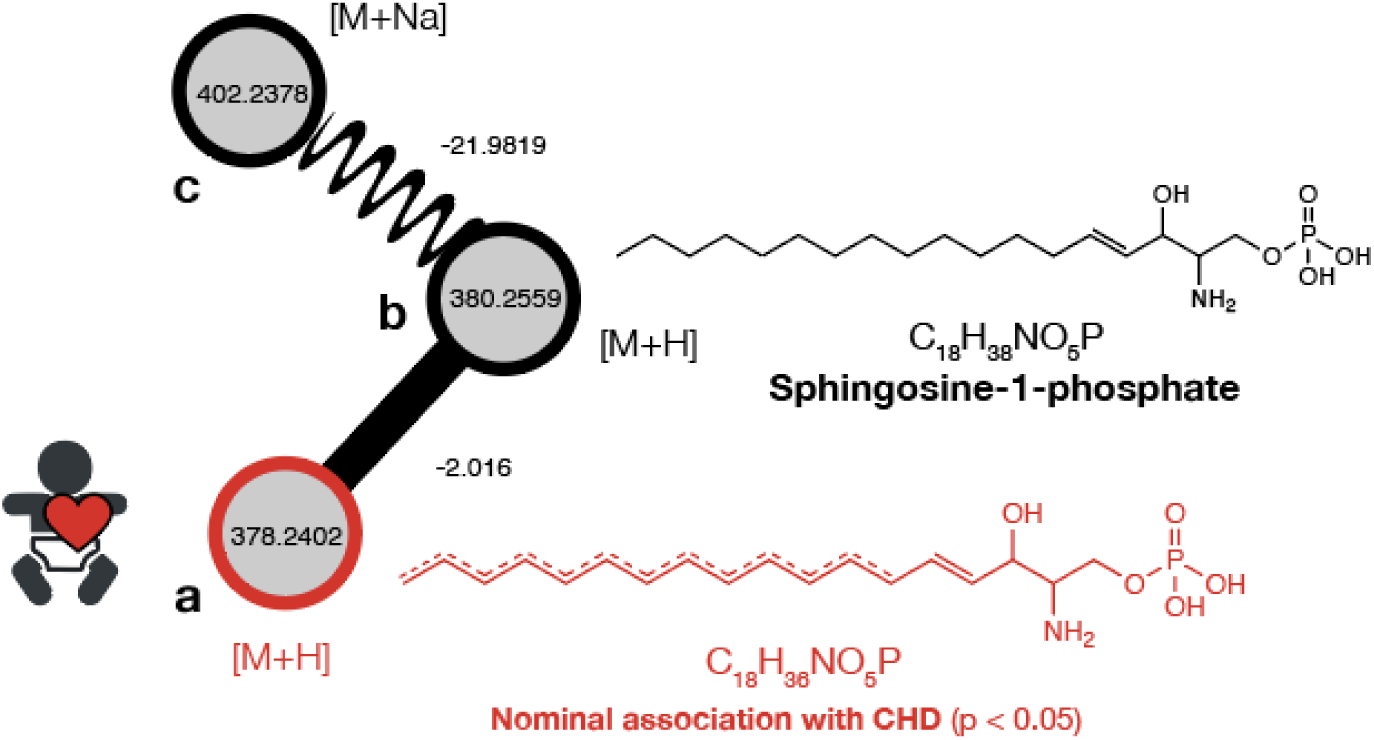
Molecular family showing a desaturated analog of S1P (a), nominally associated with CHD (p-value: 0.033) identified through mass spectral molecular networking. Nodes in the network represent MS2 (tandem mass) fingerprints of measured metabolite features, with straight lines connecting nodes with high MS2 spectral similarity (a and b) and sinewave lines connecting adducts of the same metabolite feature (b and c). The numbers on the nodes represent mass to charge ratios (*m/z*) whereas numbers on the edges represent mass to charge ratio differences.

Among the 160 CHD cases included in the study, the most common cardiac phenotype was septal defects, observed in 79 cases (49%). Other frequent phenotypes included left ventricular outflow tract obstruction (LVOTO) in 30 cases (19%), conotruncal defects in 18 cases (11%), and valve defects in 13 cases (8%). Less frequent phenotypes were atrioventricular septal defects (AVSD) with 8 cases (5%) and right ventricular outflow tract obstruction (RVOTO) with 6 cases (4%). The full distribution of phenotypes is shown in *Supplementary Table 2*. More details on the grouping can be found in Appendix A of *Oyen, et. al, 2009*^4^. We tested heterogeneity across CHD subtypes using interaction terms within CLR. No interactions were significant after FDR correction.

## Discussion

Our study is unique in focusing on the early first trimester, the critical window when the fetal heart is forming. This early gestational time point contrasts with most prior metabolomic studies of CHDs, which have analyzed maternal samples from gestational week 11 onwards, or even postpartum^14–19,24,25^, a phase where the heart continues to mature through valve and chamber remodeling, outflow tract refinement, and myocardial histodifferentiation ^26,27^.

Consequently, metabolic differences detected only later in pregnancy might reflect secondary effects of carrying a fetus with a CHD or ongoing cardiac maturation processes. Early-window sampling, as in our study, is therefore optimal for probing putative initiating maternal influences, while later sampling may capture remodeling or adaptive processes instead.

Despite targeting this key developmental window, we found no statistically significant associations between maternal metabolite levels and CHD status after FDR correction. While 69 metabolites showed nominal associations (p < 0.05), these did not withstand correction for multiple testing. These findings are consistent with those of a recent systematic review of eight maternal metabolomics studies involving 842 CHD cases, which concluded that no single metabolite has consistently differentiated CHD pregnancies from controls across studies^17^. These studies identified various metabolites spanning lipid, glucose, and amino acid pathways as potential markers, but the specific compounds differed widely between reports. Our results add to this narrative, suggesting that if maternal metabolic perturbations play a role in CHD etiology, their signals may be subtle and easily obscured by maternal physiology, study differences, random variation, inter-individual variability, or technical limitations. Moreover, the 95% Confidence Intervals (CI) around our top metabolites (*Supplementary Table 1*) allow us to rule out very large per-SD effects in early pregnancy but remain compatible with small effects.

Although no individual metabolites were statistically significantly correlated with CHD risk in our analysis, some of the annotated nominally associated metabolites are noteworthy in the context of prior work and biological plausibility. For example, one feature was annotated as a desaturated analog of S1P (*Figure 1*), a lipid signaling molecule known to regulate cardiovascular development^28^ and previously implicated in CHD pathogenesis^29,30^. In addition, recent work has highlighted a broader role for sphingolipid metabolism in cardiac biology, showing that SphK2-dependent S1P signaling is crucial for neonatal heart regeneration after injury, further supporting the relevance of this pathway in cardiac development and repair^31^. To our knowledge, this specific S1P analog has not been reported in relation to CHD, but it may point to perturbations in sphingolipid metabolism worth pursuing in future studies.

Additionally, several amino acid-related features were nominally associated with CHD (e.g., a putative arginine-related feature, the dipeptide isoleucylproline, and other amino-acid derivatives). A previous study reported negative associations between several amino acids in maternal blood and offspring CHDs, although based on samples from ∼26–28 weeks’ gestation^25^. These amino acid associations partially overlap with our nominal signals, and may inform hypotheses about amino acid metabolism and cardiac development for future research.

Finally, we detected nominal associations for additional metabolites with only limited structural information (organoheterocyclic and organic acid compounds). These have not been previously linked to CHD but could be considered in future targeted or larger-scale analyses.

### Strengths and limitations of the present study

Our study offers significant strengths relative to prior research. First, with 160 matched case–control pairs, we had a substantially larger sample size than earlier pregnancy-based metabolomic studies of CHDs, which have generally included only tens of cases rather than hundreds^20^. The absence of associations that survive FDR correction suggests that large, consistent case-control differences in early pregnancy are unlikely under our design, while smaller or subtype-specific effects cannot be excluded. If there were a large, consistent metabolic perturbation in early pregnancy among CHD mothers, that are detectable with our LC–MS/MS workflow, we would expect to observe them. However, because this platform primarily covers polar to semi-polar metabolites, we cannot exclude the possibility that stronger differences may be detectable using more lipid-focused approaches, alternative chromatographic methods, different ionization modes, or other extraction procedures.

Second, the early gestational window of our samples (weeks 4–10, when organogenesis of the heart occurs) is a distinctive strength. We captured the maternal metabolome at the biologically relevant time for CHD development, whereas studies sampling later (mid/late pregnancy) may miss early transient changes or dilute them with pregnancy’s progressing physiological alterations. Our focus on the early first trimester aligns with the period of embryonic cardiogenesis, the window when perturbations can affect formation of the heart rather than later remodeling. Third, our case-control design was rigorously matched on factors like gestational age and other potential confounders. We also applied stringent, pre-specified exclusions (*Supplementary Table 4*) to further reduce confounding. By matching and using conditional logistic regression, we minimized spurious differences due to maternal age, gestational timing, or batch effects. We also examined a broad spectrum of CHD phenotypes, rather than a single defect. This increases the overall relevance and generalizability of our findings across CHDs, but it also introduces etiologic heterogeneity that may dilute associations if metabolic perturbations are specific to particular CHD subtypes. Finally, our samples were stored under ultra-low temperature conditions (–196°C) and had not undergone any thawing prior to our metabolomics analyses. Collectively, these strengths – large sample size, early pregnancy sampling, and careful study design – position our work as a robust test of the hypothesis that maternal first-trimester metabolomic profiles differ in CHD pregnancies.

We acknowledge limitations that could have influenced our ability to detect metabolomic differences. One major consideration is the pre-analytical variability in our sample collection and handling. Unlike a dedicated biobank with uniform protocols, the first-trimester blood samples were drawn in routine care (i.e. by general practitioners) and transported (often by mail) for storage, leading to non-standardized processing times and conditions. Such random measurement noise would attenuate true case-control differences, making our results more conservative. Moreover, metabolic alterations outside our platform’s detection window may still be informative but would not appear here. Another limitation is the heterogeneity of CHD phenotypes. We performed subgroup analyses across CHD phenotypes, but these analyses were limited by smaller sample sizes in each category and did not yield significant findings. Thus, we cannot rule out that a true metabolomic signal exists for a particular CHD phenotype that we were underpowered to detect once cases were stratified. This mirrors the variation across the previous studies noted above which may partly explain why findings have been inconsistent.

## Conclusions

In conclusion, our study provides insight into the maternal metabolome during a critical window for fetal heart development and represents one of the first untargeted metabolomic investigations of CHD risk in samples collected from early in the first trimester of pregnancy. Although no metabolites survived correction for multiple testing, nominal associations within sphingolipid and amino acid pathways – including a desaturated S1P analog – offer promising leads for future hypothesis-driven research. The strengths of our study – including its relatively large sample size, carefully matched case-control design, and uniquely early gestational timing – provide a solid foundation for follow-up work. Future studies stand to benefit substantially from pre-specified pathway-focused panels, phenotype-homogeneous subgroup analyses, and independent replication, all of which would improve statistical power while reducing the multiple-testing burden. Targeted metabolomic approaches and optimized pre-analytical protocols may further enhance sensitivity to the subtle metabolic variation that could underlie CHD susceptibility in early pregnancy.

## Methods

### Identification of congenital heart defects

A CHD was identified through diagnoses recorded in either the National Patient Register (NPR)^32^ or the Cause of Death Register (CDR)^33^ within 10 years post-birth, based on designated International Classification of Diseases (ICD) codes (ICD-8: 746.0-747.4 and 759.0; ICD-10: Q20-Q26, Q8933). The classification was structured hierarchically^7,34^.

### Matching process

The Danish National Birth Cohort (DNBC) was established to investigate causal links between early-life exposures and later health outcomes, as well as to explore opportunities for disease prevention^35^. Between 1996 and 2002, the DNBC enrolled approximately 100,000 pregnancies, resulting in 96,000 live births from 92,000 unique mothers. For this study, we included women from the DNBC who provided a first-trimester blood sample, collected between gestational weeks 4 and 10. Gestational age (GA) is counted from the beginning of the last menstrual period, i.e. it runs ∼2 weeks ahead of post-conceptional age. Thus, GA weeks 4–10 correspond to ∼2–8 weeks after conception, i.e., the window of primary cardiogenesis. Cases were defined as pregnancies where the offspring was diagnosed with a CHD.

Prior to matching, pregnancies were excluded if there was history of maternal CHD, multiple offspring, offspring with a chromosomal aberration (identified through the Danish Cytogenetic Central Register) or a registration with an entry indicating a recognizable syndrome or a known cause of CHD. Pregnancies were also excluded if mothers had filled a prescription for a potentially teratogenic drug during pregnancy weeks 4-10 identified through the National Prescription Register. Finally, pregnancies resulting in offspring with a malformation due to a viral infection, such as rubella, were excluded. We also excluded pregnancies from women who had not been residing in Denmark from six months prior to conception through gestational week 10. Additional exclusions included missing data from the first DNBC interview, undergoing IVF treatment, non-cardiac malformations, and maternal pregestational diabetes, identified through diagnostic codes and/or prescriptions for antidiabetic medications filled during the six weeks prior to conception through gestational week 10. An overview of all registers used is provided at *Supplementary Table 3* and details on all the exclusion criteria are provided in *Supplementary Table 4*.

Eligible CHD cases were matched to controls based on maternal age (within one-year intervals), calendar year of sampling (within two-year intervals), and gestational week at blood draw (within one-week intervals). When multiple potential controls were available for a given case, matches were prioritized based on the smallest difference in gestational day, followed by random selection if ties remained. Matching was performed sequentially to ensure each case received its n^th^ control before proceeding to the next. Each matched pair was assigned an indicator variable encoding phenotype, case number, and status (case or control) to facilitate downstream data organization and analysis.

### Sample preparation

Maternal plasma samples were collected and sent by mail to Statens Serum Institut in Copenhagen where they were separated and stored in freezers at -196°C. Matched case-control pairs were distributed randomly over six 96-well plates (batches), made from polypropylene. To each plate pooled plasma from adult individuals, referred to as external control samples (EC) and plate specific pools of all plasma samples were added for quality control purposes. Each plate included eight EC, four process blanks (PB, extraction in empty well), four pooled samples and 80 analytical samples. On the day of extraction plasma samples were removed from the freezer and kept at room temperature until thawed. 100 µL of plasma samples were then extracted by adding 300 µL of cold methanol/acetonitrile (50:50), with subsequent shaking for 15 min at 900 rpm at room temperature, and centrifuged at 4000 rpm for 30 min at 4°C. 15 µL of extracts were then pipetted into a new 96-well plate, and evaporated under nitrogen for 1 h at 60 L/min, at room temperature.

Finally, samples were reconstituted in 100 µL reconstitution solution (consisting of 5% solvent B in 95% solvent A, see Metabolomic profiling section), shaken at 600 rpm for 15 min, and then centrifuged at 3000 rpm for 10 min at 4°C. All solvents were LCMS-grade, and were purchased from Thermo Fisher Scientific (Waltham, MA, USA). All pipetting steps were performed on a Microlab STAR automated liquid handler (Hamilton Bonaduz AG, Bonaduz, Switzerland). A Titramax 101 platform shaker (Heidolph Scientific Products GmbH, Schwabach, Germany) was used for shaking and a high-speed centrifuge 5810 R (Eppendorf, Hamburg, Germany) was used for centrifugation.

### Metabolomics profiling

Plasma samples were submitted to untargeted metabolomics profiling using LC-MS/MS at Statens Serum Institut, Copenhagen, Denmark between November and December 2022. The metabolite profiling was performed using a LC-MS/MS platform consisting of a timsTOF Pro mass spectrometer with an VIP-HESI ion-source for electrospray ionization, Bruker Daltonics (Billerica, MA, US) coupled to a UHPLC Elute LC system, Bruker Daltonics (Billerica, MA, US). The chromatographic separation system included a binary pump, an autosampler with cooling function, and a column oven with temperature control. Infusion of reference solution, used for external and internal mass calibration, was performed using an additional isocratic pump, Azura Pump P4.1S (Knauer, Berlin, Germany). The chromatographic separation was performed on an Acquity HSS T3 (100 Å, 2.1 mm x 100 mm, 1.8 µm) column (Waters, Milford, MA, US). The mobile phase consisted of solvent A (99.8% water and 0.2% formic acid) and B (49.9% methanol, 49.9% acetonitrile and 0.2% formic acid). The initial chromatographic conditions were 99% mobile phase A, which was kept for 1.5 min, followed by a linear gradient to 95% mobile phase B during 8.5 min, isocratic conditions at 95% mobile phase B for 2.5 min before going back to 99% mobile phase A for 2.4 min. Total run time for each injection was 15 min and the analysis time for a full 96-well plate was approximately 25 h. Samples were maintained at +15°C in the autosampler, 5 µL were loaded to the column with a flow rate of 0.4 mL/min and a column temperature of 40°C. Mass spectrometric analysis was performed in Q-TOF mode with TIMS off, and auto MS/MS on using the following settings: ionization mode set to positive ionization, mass range set to 20–1100 *m/z*, a spectral rate of 9 Hz (MS1 sampling time 0.11s) and a cycle time of 0.5 s. The source settings were set as, capillary voltage: 4500 V, nebulizer gas: 2.2 Bar, dry gas flow: 10 l/min, dry gas temperature: 220°C. Tune settings as follows: funnel 1 RF and funnel 2 RF: 200 Vpp, isCID: 0 eV, multipole RF: 60 Vpp, deflection Delta: 60 V, quadrupole ion energy: 5 eV with a low mass set to 60 *m/z*, Collision cell energy set to 7 eV with a pre pulse storage of 5 µs. Stepping was used in basic mode with a collision RF from 250 to 750 Vpp, transfer time 20−50 μs and timing set to 50% for both. For MS/MS, the collision energy ranged from 100% to 250% relative to the base collision energy of 7 eV with timing set to 50% for both. Auto MS/MS was applied with active exclusion after 3 spectra and the release time set to 0.15 min. Dynamic MS/MS spectra acquisition was performed using a target intensity of 20,000 counts, max MS/MS spectra acquisition of 30 Hz (0.03 sec) and min MS/MS spectra acquisition of 16 Hz (0.06 sec). A precursor exclusion list was used with an exclusion of mass range of 20−60 *m/z*. Sodium formate clusters were applied for instrument mass calibration and for internal recalibration of individual samples. A Precursor Exclusion list was used with Exclusion of mass range of 20−60. For system suitability and quality control check see *Supplementary Note*.

### Metabolomics preprocessing

Bruker .d files were exported to the .mzML format using Bruker’s DataAnalysis software and subsequently preprocessed using the Ion Identity Network workflow in MZmine (version 3.9.0)^36–38^. Data were imported and centroid mass lists created using the advanced import option with MS1 intensity above 2.5E2 and MS2 intensity above 0 retained. The chromatogram was built through the ADAP Chromatogram Builder by using the following parameters: minimum consecutive scans: 5, minimum intensity for consecutive scans: 2.5E2, minimum absolute height: 1E3, and *m/z* tolerance: 0.002 *m/z* or 5 ppm. A scan filter was applied within the ADAP Chromatogram Builder with chromatogram retention time 0.3 to 13 min. retained for MS1. The chromatograms were further smoothed using the Loess smoothing algorithm with a retention time width (scans) of 5. Features were deconvoluted using the Local Minimum Feature Resolver with MS/MS scan pairing set to: MS1 to MS2 precursor tolerance: 0.001 *m/z* or 3 ppm, retention time filter: use feature edges, minimum relative feature height: 25%, minimum required signals: 1, and further parameters set to: dimension: retention time, chromatography threshold: 85%, minimum search range retention time: 0.01, minimum relative height: 0%, minimum absolute height: 1E3, min ratio of peak top/edge: 3, peak duration range: 0.02−0.4 min, minimum scans: 5. Peaks were deisotoped by using the 13C isotope filter, with parameters set to: *m/z* tolerance (intra-sample): 0.002 *m/z* or 5 ppm, retention time tolerance: 0.1 min, monotonic shape: on, maximum charge: 2, representative isotope: most intense, never remove feature with MS2: on. Peaks from all samples were aligned using the Join Aligner with parameters set to: *m/z* tolerance (sample-to-sample): 0.002 *m/z or* 5 ppm, weight for *m/z*: 5, retention time tolerance: 0.1 min, weight for RT: 1. Gap-filling was performed using the Peak Finder (multithreaded) with parameters set to: intensity tolerance: 20%, *m/z* tolerance: 0.001 *m/z* or 5 ppm, retention time tolerance 0.1 min and minimum scans (data points): 1. The Duplicate Peak Filter was used to remove redundancy with parameters set to: filter mode: new average, *m/z* tolerance: 0.001 *m/z* or 5ppm, retention time tolerance: 0.03 min. The metaCorrelate function was used to find correlating peak shapes with parameters set to: retention time tolerance: 0.1 min, minimum feature height: 1E3, intensity threshold for correlation: 2.5E3, min samples in all: 2 (abs), min samples in group: 0 (abs), min %−intensity overlap: 60%, exclude gap-filled features: on. Parameters for the feature shape correlation grouping were set as follows, min data points: 5, min data points on edge: 2, measure: Pearson, min feature shape correlation: 85%. Ion identity networking parameters were set to *m/z* tolerance: 0.002 *m/z* or 5 ppm, check: one feature, min height: 1E3 with ion identity library parameters set to: MS mode: positive, maximum charge: 2, maximum molecules/cluster: 2, adducts: M+H, M+Na, M+K, modifications: M-H2O, M-NH3. Further ion identity networks were added with *m/z* tolerance: 0.002 *m/z* or 5 ppm, min height: 1E3 and ion identity library parameters set to MS mode: positive, maximum charge: 2, maximum molecules/cluster: 6, adducts: M+H, M+Na, M+NH4, modifications: M-H2O, M-NH3, M-2H2O, M-3H2O, M-4H2O, M-5H2O and *m/z* tolerance: 0.002 *m/z* or 5 ppm, min height: 1E3, and annotation refinement on with parameters set to delete smaller networks: link threshold: 4, delete networks without monomer: on, and ion identity library parameters set to MS mode: positive, maximum charge: 2, maximum molecules/cluster: 2, adducts: M+H, M+Na, M+K, modifications: M-H2O, M-NH3. Finally, two feature tables were exported in the .csv format. One feature table containing all extracted mass spectral features and another feature table filtered for mass spectral features with associated fragmentation spectra (MS2). An aggregated list of MS2 fragmentation spectra was exported in the .mgf format and submitted to ion identity feature-based mass spectral molecular networking through the Global Natural Products Social Molecular Networking Platform (GNPS)^37,39^.

For downstream statistical analysis the feature table containing all mass spectral features was used. First, we removed samples, where EDTA was not detected during the metabolomics experiments, suggesting that a different anticoagulant had been used during blood collection. Anticoagulants other than EDTA can introduce noise in mass spectrometric data and have been shown to affect the metabolite profile in ways that are not easy to adjust for^40^. An overview of sample selection is shown in *Supplementary Figure 4*.

Next, to reduce redundancy, connected ion adducts were merged. The adduct observed in the greatest number of samples and with the highest median signal intensity was chosen. Further all mass spectral feature signals with a relative intensity less than 5 times the mean relative intensity in all paper blank samples and metabolite features present in fewer than 75% of the samples were removed. To reduce technical variation, we furthermore filtered out metabolites, which showed a relative standard deviation >30% across repeated injections of external controls (EC) and plate-specific pooled samples (PO). In a subsequent data-cleaning step, we excluded a molecular family of features considered likely MS contaminants, based on their very early elution (<0.6 min) and annotation consistent with MS contaminant sodium formate cluster ions. Missing features were imputed by choosing a random number between 0 and the lowest detected value for each feature. The overall sparsity of the feature table prior to imputation was 3.6%. To mitigate batch effects, we used the WavelCA 2.0 algorithm ^41^ and centered and univariance scaled each metabolite, converting outliers deviating more than 5 standard deviation units from the mean to the values 5 and -5 respectively. The final dataset consisted of a total of 320 samples and 1,471 metabolite features measured.

### Metabolite identification

Metabolite annotation was performed using mass spectral molecular networking through the GNPS Platform^37,39,42^, whereafter MS2LDA substructure information^43^ was incorporated through the ^44^ MolNetEnhancer workflow^45^ . Finally, we retrieved *in silico* structure annotation through Sirius+CSI:FingerID^46,47^ and CANOPUS^44^. The metabolite feature annotation confidence level was defined according to the Metabolites Standard Initiative^48^, as level 2 (plausible 2D structure), level 3 (plausible metabolite class) or level 4 (unknown). Metabolite annotations for nominally significant features can be found in *Supplementary Table 1*. A detailed description of GNPS mass spectral molecular networking parameters can be found in the *Supplementary Note*.

### Statistical Analysis

Metabolite intensities were standardized using Z-score transformation, ensuring comparability of effect sizes across metabolites and a stable variance for regression. We evaluated the association between each metabolite and CHD case-control status using univariate conditional logistic regression models^49^. Each model included a single metabolite as the exposure and was stratified by the matched pair identifier to account for the 1:1 matching. Multivariable conditional logistic regression models, adjusting for risk factors for CHD that could also potentially influence maternal metabolite profiles were also developed. These covariates included the sex of the child (binary), maternal pre-pregnancy BMI (continuous), gestational age at sampling (continuous), maternal smoking prior to pregnancy onset (categorical: non-smoker, occasional, daily <15 cigarettes, daily ≥15 cigarettes), maternal alcohol use prior to pregnancy onset (ordered categories from 0 to 5 units/week in 0.5-unit increments), and parity (categorical). For each metabolite, the model estimated an odds ratio (OR) and corresponding *p*-value, representing the association between a one-standard-deviation increase in the metabolite and the odds of CHD in the offspring. Missing values in covariates were imputed using chained equations (`mice` package), without imputing the outcome or matching variables.

To investigate whether metabolite–CHD associations differed across CHD subtypes, we conducted an interaction analysis within the same CLR framework. For each metabolite, we fitted two models: (i) a main-effects model including the Z-scored metabolite and CHD subtype (defined from the case’s primary diagnosis; subtypes with ≤5 cases (APVR, Other specified defects) were combined into a single “Other” category), and (ii) an interaction model including an additional metabolite × CHD subtype interaction term. Both models were stratified by matched pair and adjusted for the same covariates as above. A likelihood ratio test (LRT) comparing the interaction model to the main-effects model provided a global p-value for heterogeneity of the metabolite–CHD association across subtypes. Subtype-specific ORs (per 1-SD increase in the metabolite) were derived from the interaction model by combining the relevant regression coefficients.

Given the large number of metabolites tested, we applied a false discovery rate control to limit false positives. For the case–control analyses, FDR correction, using the Benjamini–Hochberg method^50^, was applied to the per-metabolite p-values from the CLR models; for the interaction analyses, FDR correction was applied to the global interaction p-values from the LRTs. Metabolites with an FDR-adjusted *p*-value below 0.05 were considered statistically significant. All analyses were conducted in R (version 4.3). Conditional logistic regression and interaction analyses were implemented using the *clogit* function from the *survival* package^51^.

### Ethical approval and consent to participate

The DNBC was approved by the Danish Data Protection Agency and the Committee on Health Research Ethics (case no. (KF) 01-471/94). The women participating in the DNBC provided written informed consent at the time of their enrolment in the cohort. The protocol for this study was approved by the Regional Scientific Ethical Committee of Copenhagen (H-16043279) and by the DNBC scientific management group and steering committee. Data handling in this study was approved by Statens Serum Institut (SSI) and is covered by the general approval for research at SSI given by the Danish Data Protection Agency. Participants or the public had no involvement in how the study was designed, conducted, reported, and disseminated.

## Data Availability

The individual-level data underlying the results cannot be made publicly available due to data protection regulations. However, access to the data for researchers may be approved with some restrictions if necessary conditions are met. Data is part of the Danish National Birth Cohort (DNBC) data collection and can be requested for research purposes. More information regarding access to data and application procedures can be found on the DNBC website http://www.dnbc.dk/access-to-dnbc-data.

## Acknowledgements

The Danish National Birth Cohort was established with a significant grant from the Danish National Research Foundation. Additional support was obtained from the Danish Regional Committees, the Pharmacy Foundation, the Egmont Foundation, the March of Dimes Birth Defects Foundation, the Health Foundation and other minor grants. The DNBC Biobank is part of the Danish National Biobank resource and has been supported by the Novo Nordisk Foundation and the Lundbeck Foundation. The project was supported by grants from Børnehjertefonden (16-R99-A5073-26037) and the Novo Nordisk Foundation (NNF16OC0020968).

## Author contributions

KN and FO contributed equally to this work. KN and FO performed the statistical analyses under the supervision of JW, GC, FG and BF. FO and ME carried out metabolomics preprocessing and chemical structural annotation and contributed to the interpretation of the metabolomics results. SGB and NM developed the LC–MS/MS method, generated the metabolomics data and performed initial quality control. ME supervised metabolomics quality control, data acquisition, preprocessing, chemical structural annotation and downstream analyses. ABS, ML, and MM conceived the study within the DNBC and ABS, GC, and ML acquired register and questionnaire data. ABS, MM, and BF obtained approvals and funding for the study. BF provided overall supervision of the study. KN wrote the first draft of the manuscript with contributions from ME. All authors contributed to the interpretation of the results and reviewed, edited and approved the final manuscript.

## Data and code availability

The code underlying this article is provided via GitHub: https://github.com/ssi-dk/CHD-metabolomics.

