## Supplementary for "Early pregnancy metabolomics and risk of offspring heart defects: a matched case-control study"

* equal contribution

### Supplementary Note

#### System suitability and quality control

Quality control procedures for the metabolite profiling were divided into three main categories as described previously: system suitability test (SST), batch evaluation and project evaluation[1]. In brief, during SST, mass spectral and chromatographic performance was evaluated prior to each batch by injecting 5 µL of standard sample A and B. Standard sample A consisted of leucine enkephalin (1.8 µM in water:acetonitrile, 50:50) and standard sample B consisted of a labelled amino acid and acylcarnitine mixture (Cambridge Isotope Laboratories, Tewksbury, MA, USA) dissolved in methanol and reconstitution solution (5% solvent B in solvent A). System suitability was then evaluated based on retention time (RT) deviation (< 0.2 minutes), mass accuracy (< 2 ppm) and relative standard deviation (< 20%) of extracted peak intensities for both samples A and B, as well as MS/MS spectral similarity for sample B. Batch evaluation was performed by evaluating mass spectral and chromatographic performance through retention time deviation (<0.2 min), mass accuracy (<2 ppm) and coefficient of variation (<20%) of sixteen quality control metabolites (= metabolites typically found in high abundance in these sample types) in pooled, EC and PB samples. Furthermore, we controlled for potential carry-over by ensuring that quality control metabolites were not present in the paper blank samples. Feature extraction for the SST and batch evaluation was performed in Metaboscape (Bruker, Billerica, MA, United States).

#### Mass spectral molecular networking parameters

A mass spectral molecular network was created through the Global Natural Products Social Molecular Networking Platform (GNPS)[2] (http://gnps.ucsd.edu) using the feature-based molecular networking workflow[3]. The data was filtered by removing all MS/MS fragment ions within +/- 17 Da of the precursor *m/z*. MS/MS spectra were window filtered by choosing only the top 6 fragment ions in the +/- 50 Da window throughout the spectrum. The precursor ion mass tolerance was set to 0.02 Da and the MS/MS fragment ion tolerance to 0.02 Da. A molecular network was then created where edges were filtered to have a cosine score above 0.7 and more than 4 matched peaks. Further, edges between two nodes were kept in the network if and only if each of the nodes appeared in each other's respective top 10 most similar nodes. Finally, the maximum size of a molecular family was set to 100, and the lowest scoring edges were removed from molecular families until the molecular family size was below this threshold. The spectra in the network were then searched against GNPS spectral libraries. The library spectra were filtered in the same manner as the input data. All matches kept between network spectra and library spectra were required to have a score above 0.7 and at least 4 matched peaks. The molecular networks were visualized using Cytoscape software[4].

**Supplementary Table 1. Annotations of significant metabolites from adjusted Conditional Logistic Regression (CLR).** Odds Ratios (OR) from CLR reported with unit = 1 SD increase in metabolite.

| Feature ID | Case vs control (per 1 SD) OR [95% CI] | p-value | FDR | parent mass | RT mean | MS level | Curated Annotation | Annotation Method | Metabolite Annotation Level |
|---|---|---|---|---|---|---|---|---|---|
| 525 | 1.71 (1.25, 2.32) | 0.001 | 0.61 | 418.8791 | 0.5473 | MS2 | - | - | 4 |
| 527 | 1.66 (1.23, 2.23) | 0.001 | 0.61 | 486.8665 | 0.5474 | MS1 | - | - | 4 |
| 504 | 1.58 (1.18, 2.11) | 0.002 | 0.78 | 493.8589 | 0.5258 | MS1 | - | - | 4 |
| 421 | 1.54 (1.17, 2.02) | 0.002 | 0.78 | 437.8864 | 0.5253 | MS1 | - | - | 4 |
| 500 | 1.58 (1.17, 2.14) | 0.003 | 0.78 | 380.8969 | 0.5277 | MS2 | Amino acids and derivatives | GNPS | 3 |
| 482 | 1.55 (1.15, 2.08) | 0.004 | 0.89 | 432.8564 | 0.5223 | MS1 | - | - | 4 |
| 352 | 1.52 (1.14, 2.05) | 0.005 | 0.91 | 428.8823 | 0.5247 | MS1 | - | - | 4 |
| 524 | 1.57 (1.14, 2.16) | 0.005 | 0.91 | 356.9085 | 0.5513 | MS1 | - | - | 4 |
| 2354 | 1.46 (1.11, 1.91) | 0.007 | 0.91 | 229.1543 | 1.4653 | MS2 | Isoleucylproline | Sirius+CSI:FingerID | 2–3 |
| 606 | 1.47 (1.11, 1.95) | 0.007 | 0.91 | 828.8058 | 0.5848 | MS1 | - | - | 4 |
| 1440 | 1.43 (1.10, 1.86) | 0.007 | 0.91 | 381.9859 | 0.8645 | MS1 | - | - | 4 |
| 1827 | 0.67 (0.50, 0.90) | 0.008 | 0.91 | 279.9876 | 1.1295 | MS1 | - | - | 4 |
| 612 | 1.52 (1.11, 2.07) | 0.009 | 0.91 | 567.8916 | 0.578 | MS1 | - | - | 4 |
| 1708 | 1.38 (1.08, 1.77) | 0.010 | 0.91 | 298.0249 | 1.0226 | MS1 | - | - | 4 |
| 675 | 0.68 (0.51, 0.91) | 0.010 | 0.91 | 124.9227 | 0.5899 | MS1 | - | - | 4 |
| 448 | 1.52 (1.10, 2.10) | 0.011 | 0.91 | 474.8334 | 0.5249 | MS1 | - | - | 4 |
| 638 | 1.48 (1.09, 2.01) | 0.012 | 0.91 | 854.8113 | 0.5835 | MS1 | - | - | 4 |

| Feature ID | Case vs control (per 1 SD) OR [95% CI] | p-value | FDR | parent mass | RT mean | MS level | Curated Annotation | Annotation Method | Metabolite Annotation Level |
|---|---|---|---|---|---|---|---|---|---|
| 650 | 0.69 (0.52, 0.92) | 0.012 | 0.91 | 818.7719 | 0.5887 | MS1 | - | - | 4 |
| 491 | 1.44 (1.08, 1.91) | 0.012 | 0.91 | 296.9439 | 0.5281 | MS1 | - | - | 4 |
| 428 | 1.46 (1.08, 1.96) | 0.012 | 0.91 | 492.844 | 0.5252 | MS1 | - | - | 4 |
| 1079 | 1.41 (1.06, 1.86) | 0.017 | 0.96 | 370.7413 | 0.6548 | MS1 | - | - | 4 |
| 442 | 1.43 (1.07, 1.93) | 0.017 | 0.96 | 232.9279 | 0.5211 | MS2 | Amino acids and derivatives | GNPS | 3 |
| 1914 | 0.73 (0.56, 0.95) | 0.018 | 0.96 | 236.9908 | 1.1269 | MS1 | - | - | 4 |
| 626 | 0.71 (0.53, 0.94) | 0.018 | 0.96 | 328.8851 | 0.5881 | MS1 | - | - | 4 |
| 685 | 0.70 (0.52, 0.94) | 0.018 | 0.96 | 614.81 | 0.5895 | MS1 | - | - | 4 |
| 644 | 0.70 (0.52, 0.94) | 0.018 | 0.96 | 38.9632 | 0.5863 | MS1 | - | - | 4 |
| 487 | 1.41 (1.05, 1.89) | 0.021 | 0.96 | 255.943 | 0.5219 | MS1 | - | - | 4 |
| 548 | 1.40 (1.05, 1.86) | 0.021 | 0.96 | 492.8835 | 0.5569 | MS1 | - | - | 4 |
| 651 | 0.71 (0.53, 0.95) | 0.023 | 0.96 | 547.8263 | 0.5888 | MS1 | - | - | 4 |
| 583 | 1.44 (1.05, 1.96) | 0.023 | 0.96 | 431.9169 | 0.5762 | MS1 | - | - | 4 |
| 684 | 0.71 (0.53, 0.96) | 0.024 | 0.96 | 546.8227 | 0.5896 | MS1 | - | - | 4 |
| 2223 | 0.71 (0.53, 0.96) | 0.024 | 0.96 | 259.0047 | 1.3435 | MS2 | Organic acids and derivatives | Sirius+CSI:FingerID | 3 |
| 461 | 1.35 (1.04, 1.76) | 0.025 | 0.96 | 254.932 | 0.5237 | MS1 | - | - | 4 |
| 10866 | 0.75 (0.59, 0.97) | 0.026 | 0.96 | 581.2951 | 10.6535 | MS1 | - | - | 4 |
| 599 | 1.41 (1.04, 1.92) | 0.026 | 0.96 | 90.9766 | 0.5799 | MS2 | Arginine related metabolite | GNPS | 2–3 |
| 359 | 1.41 (1.04, 1.92) | 0.027 | 0.96 | 494.8747 | 0.5254 | MS1 | - | - | 4 |

| Feature ID | Case vs control (per 1 SD) OR [95% CI] | p-value | FDR | parent mass | RT mean | MS level | Curated Annotation | Annotation Method | Metabolite Annotation Level |
|---|---|---|---|---|---|---|---|---|---|
| 1084 | 1.34 (1.03, 1.73) | 0.027 | 0.96 | 200.8591 | 0.6186 | MS1 | - | - | 4 |
| 672 | 0.72 (0.54, 0.97) | 0.028 | 0.96 | 532.8478 | 0.5878 | MS1 | - | - | 4 |
| 6786 | 1.36 (1.03, 1.79) | 0.030 | 0.96 | 386.2901 | 8.3131 | MS1 | - | - | 4 |
| 7193 | 1.37 (1.03, 1.83) | 0.031 | 0.96 | 371.2985 | 8.823 | MS1 | - | - | 4 |
| 688 | 0.73 (0.55, 0.97) | 0.032 | 0.96 | 616.809 | 0.5898 | MS1 | - | - | 4 |
| 478 | 1.54 (1.03, 2.30) | 0.034 | 0.96 | 67.5101 | 0.5225 | MS1 | - | - | 4 |
| 8064 | 1.32 (1.02, 1.72) | 0.035 | 0.96 | 378.2402 | 9.5461 | MS2 | Desaturated analog of sphingosine-1-phosphate | GNPS, Sirius+CSI:FingerID | 2–3 |
| 1568 | 0.75 (0.58, 0.98) | 0.035 | 0.96 | 208.9972 | 0.9028 | MS2 | Organoheterocyclic compound | CANOPUS | 3 |
| 10762 | 1.31 (1.02, 1.69) | 0.036 | 0.96 | 521.3313 | 10.6418 | MS1 | - | - | 4 |
| 635 | 1.35 (1.02, 1.78) | 0.036 | 0.96 | 760.8189 | 0.5858 | MS1 | - | - | 4 |
| 2034 | 1.31 (1.02, 1.68) | 0.037 | 0.96 | 193.0019 | 1.167 | MS2 | Organoheterocyclic compound | - | 3 |
| 2081 | 1.37 (1.02, 1.84) | 0.037 | 0.96 | 166.0858 | 1.2161 | MS1 | - | - | 4 |
| 958 | 1.33 (1.02, 1.73) | 0.037 | 0.96 | 119.0894 | 0.6707 | MS1 | - | - | 4 |
| 419 | 0.76 (0.58, 0.98) | 0.037 | 0.96 | 241.9995 | 0.5239 | MS1 | - | - | 4 |
| 666 | 1.39 (1.02, 1.91) | 0.037 | 0.96 | 447.8909 | 0.5841 | MS1 | - | - | 4 |
| 569 | 0.77 (0.60, 0.99) | 0.038 | 0.96 | 207.0505 | 0.5633 | MS1 | - | - | 4 |
| 1195 | 1.34 (1.02, 1.78) | 0.038 | 0.96 | 83.0602 | 0.7179 | MS1 | - | - | 4 |
| 623 | 0.74 (0.55, 0.98) | 0.039 | 0.96 | 682.7973 | 0.5894 | MS1 | - | - | 4 |
| 13985 | 0.77 (0.60, 0.99) | 0.039 | 0.96 | 301.2109 | 11.6206 | MS1 | - | - | 4 |

| Feature ID | Case vs control (per 1 SD) OR [95% CI] | p-value | FDR | parent mass | RT mean | MS level | Curated Annotation | Annotation Method | Metabolite Annotation Level |
|---|---|---|---|---|---|---|---|---|---|
| 654 | 1.34 (1.01, 1.78) | 0.039 | 0.96 | 515.8782 | 0.5844 | MS1 | - | - | 4 |
| 690 | 0.74 (0.55, 0.99) | 0.040 | 0.96 | 480.834 | 0.592 | MS1 | - | - | 4 |
| 1045 | 1.36 (1.01, 1.82) | 0.041 | 0.96 | 258.8176 | 0.6172 | MS1 | - | - | 4 |
| 631 | 1.34 (1.01, 1.79) | 0.043 | 0.96 | 719.8397 | 0.5829 | MS1 | - | - | 4 |
| 581 | 1.36 (1.01, 1.84) | 0.043 | 0.96 | 363.9295 | 0.5774 | MS1 | - | - | 4 |
| 7882 | 1.34 (1.01, 1.77) | 0.043 | 0.96 | 473.3067 | 9.4031 | MS1 | - | - | 4 |
| 591 | 1.36 (1.01, 1.84) | 0.045 | 0.96 | 227.9548 | 0.5743 | MS1 | - | - | 4 |
| 691 | 0.74 (0.55, 0.99) | 0.046 | 0.96 | 630.7841 | 0.5909 | MS1 | - | - | 4 |
| 621 | 1.36 (1.01, 1.83) | 0.046 | 0.96 | 812.8317 | 0.5777 | MS1 | - | - | 4 |
| 870 | 1.37 (1.01, 1.88) | 0.047 | 0.96 | 360.0748 | 0.6576 | MS1 | - | - | 4 |
| 1419 | 1.33 (1.00, 1.76) | 0.047 | 0.96 | 317.0564 | 0.8407 | MS2 | Amino acid and derivatives | Sirius+CSI:FingerID | 3 |
| 646 | 1.33 (1.00, 1.77) | 0.048 | 0.96 | 584.8622 | 0.5842 | MS1 | - | - | 4 |
| 686 | 0.75 (0.57, 1.00) | 0.048 | 0.96 | 190.9118 | 0.5906 | MS1 | - | - | 4 |
| 628 | 1.35 (1.00, 1.83) | 0.049 | 0.96 | 846.8251 | 0.5782 | MS1 | - | - | 4 |

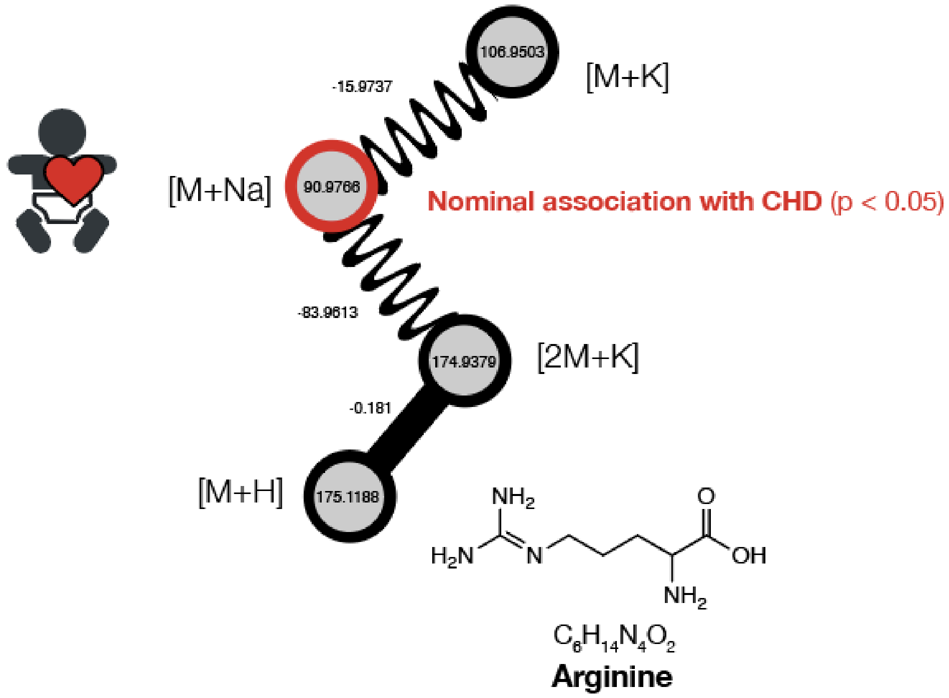


**Supplementary Figure 1**. Molecular family showing a possible unannotated structural analogue of arginine, nominally associated with CHD (p-value: 0.031) identified through mass spectral molecular networking. Nodes in the network represent MS2 fingerprints of measured metabolite features, with straight lines connecting nodes with high MS2 spectral similarity and sinewave lines connecting adducts of the same metabolite feature. The numbers on the nodes represent mass to charge ratios (*m/z*) whereas numbers on the edges represent mass to charge ratio differences.

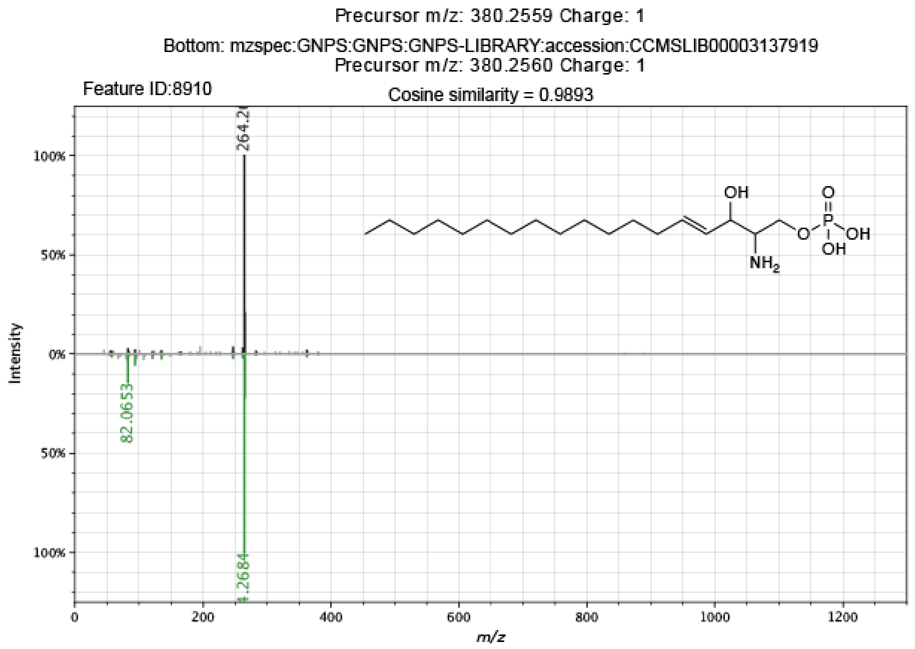


**Supplementary Figure 2**. Spectral mirror plot of mass spectral feature with a spectral library match to sphingosine-1-phosphate. The top panel shows the query spectrum whereas the bottom panel shows the library spectrum from GNPS. The mirror plot was generated using the Metabolomics Spectrum Resolver Web Service[5].

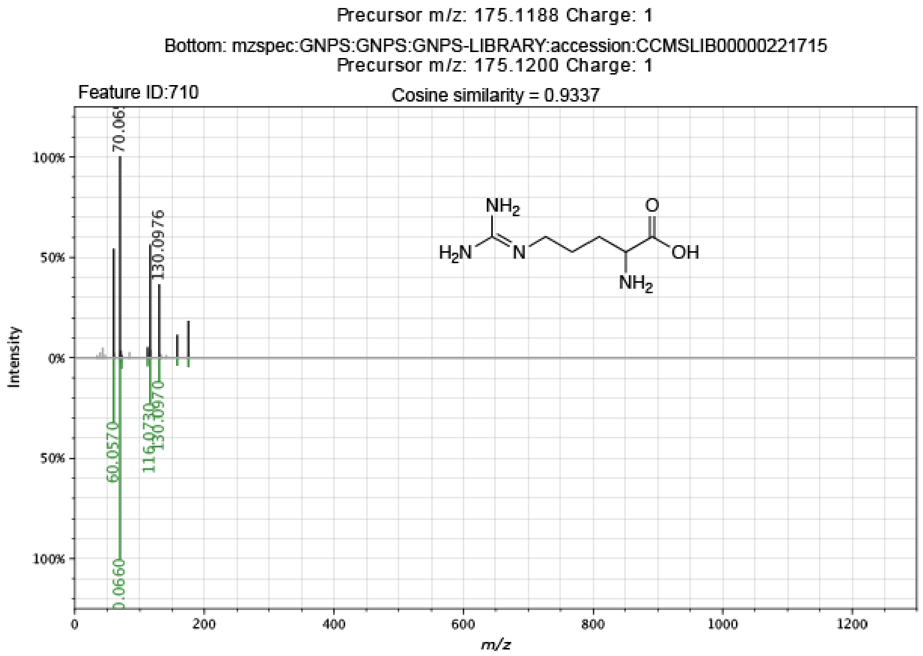


**Supplementary Figure 3**. Spectral mirror plot of mass spectral feature with a spectral library match to arginine. The top panel shows the query spectrum whereas the bottom panel shows the library spectrum from GNPS. The mirror plot was generated using the Metabolomics Spectrum Resolver Web Service[5].

**Supplementary Table 2**. **CHD phenotypes.** Bold text denotes the phenotypes used in all analyses in the paper.

| CHD phenotype | N |
|---|---|
| **Conotruncal defects** | |
| tetralogy of Fallot (TOF) | 11 |
| other conotruncal defects | <5 |
| double outlet right ventricle (DORV) | <5 |
| d-transposition of great arteries (TGA) | <5 |
| truncus arteriosus | <5 |
| **Atrioventricular septal defect (AVSD)** | 8 |
| **Anomalous pulmonary venous return (APVR)** | <5 |
| **Left Ventricular Outflow Tract Obstruction (LVOTO)** | |
| coarctation of aorta with intact ventricular septum (COA) | 13 |
| hypoplastic left heart syndrome (HLHS) | 9 |
| valvar aortic stenosis (VAS) | 7 |
| ventricular myocardial shortening (VMS_A) | <5 |
| **Right Ventricular Outflow Tract Obstruction (RVOTO)** | |
| valve pulmonary stenosis (VPS) | 6 |
| **Septal defects** | |
| ventricular septal defect (VSD) | 51 |
| atrial septal defect (ASD) | 18 |
| ASD & VSD | 7 |
| unspecified septal defects | <5 |
| **Valve defects** | 13 |
| **Other specified** (e.g. arterial and venous malformation and other specified malformations not included in other groups) | 5 |

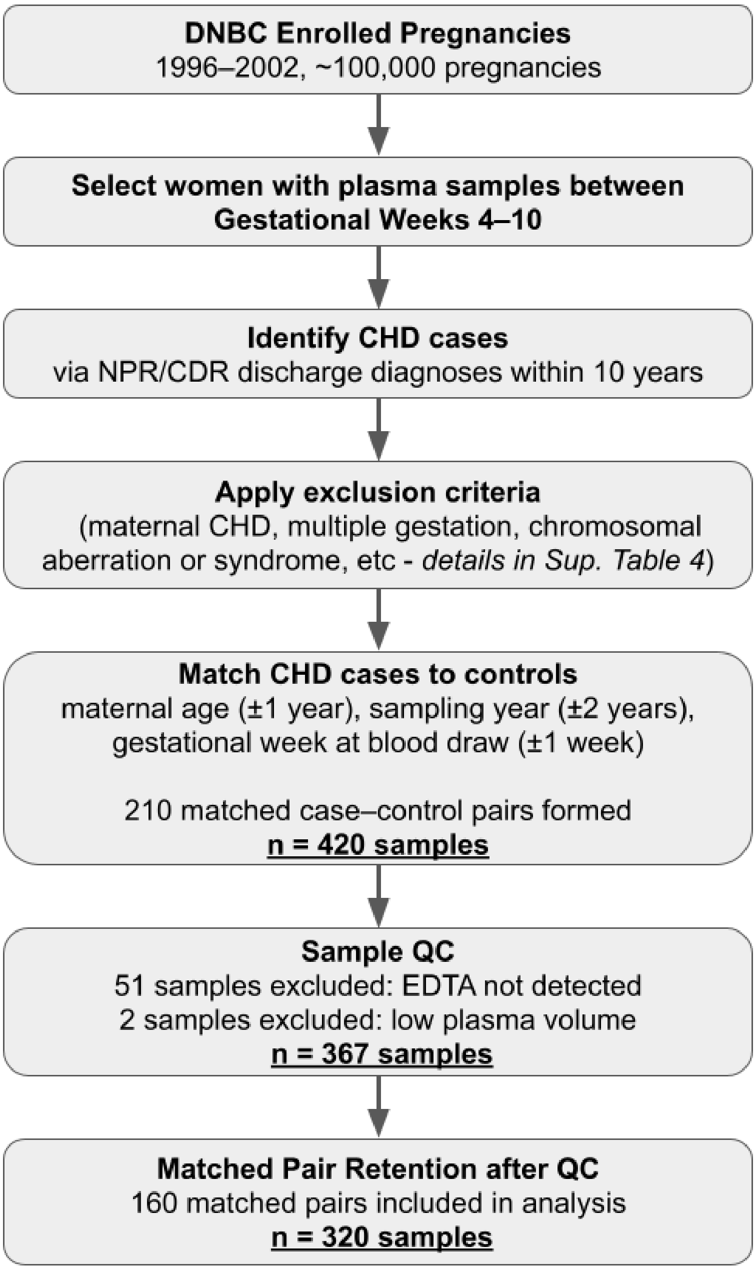


**Supplementary Figure 4. Flowchart of sample selection, matching, and QC.** Participants were drawn from DNBC and included in the study if an early first-trimester blood plasma sample was available (gestational weeks 4–10). CHD cases were identified from registry data using ICD codes and matched to controls based on maternal age, sampling year, and gestational week, with samples excluded based on the criteria detailed in Supplementary Table 4. After matching, 420 samples were selected. Metabolomics analyses suggested that in 53 samples, an anticoagulant different from EDTA was used and after removing unmatched individuals, 160 complete case–control pairs (320 samples) remained for statistical analysis.

**Supplementary Table 3. Overview of the Danish Health Registers Used in the Study.**

| Register[a] | Abbreviation | Time period | Contains information on | Reference |
|---|---|---|---|---|
| The Civil Registration System | CRS | 1968- | All individuals who have had residency in Denmark or Greenland. Contains covariates such as PIN-number, address, civil status and kinship. | [6] |
| The National Patient Register | NPR | 1977- | Contacts to Danish hospitals including diseases (ICD-codes), treatment, date of admission and discharge. | [7] |
| The Medical Birth Register | MBR | 1973- | The course and result of births, complications during pregnancy and delivery. | [8] |
| The Danish Register of Causes of Death | CDR | 1970- | Date of death, results from autopsies, cause of death and contributing causes of death retrieved from death certificates. | [9] |
| The Danish Cytogenetic Central Register | DCCR | 1960- | Postnatal or prenatal chromosome examinations, molecular genetic examinations and biochemical examinations. | https://www.fagperson.auh.dk/afdelinger/klinisk-genetisk-afdeling/dccr |
| The National Prescription Register | DNPR | 1994- (complete since 1995) | All filled (redeemed) prescriptions in Danish pharmacies. Information on package size, dosage, substitution, price, location. | [10] |
| The Danish National IVF-Register | IVFR | [illegible]94- | All treatments with in vitro fertilization (IVF), intracytoplasmic sperm injection, frozen embryo replacements, inseminations and egg donations. | [11] |

Abbreviations: PIN: Personal Identification Number, ICD: International Classification of Diseases, IVF: In Vitro Fertilization

[a]All Danish registers can be linked on individual-level by use of the Personal Identification Number

**Supplementary Table 4. Definitions of the Exclusion Criteria Used in the Study.**

| Exclusion criterion | Requirement | | Data source |
|---|---|---|---|
| Residency in Denmark during pregnancy | Mothers not living in Denmark for at least 6 months before conception and until GW10 | | CRS |
| Maternal congenital heart defect | Mother with ≥1 diagnosis of congenital heart defect (as defined for outcome) during lifetime | | NPR, CDR |
| Not singleton pregnancy | Children from multiple births | | MBR |
| Chromosomal aberration | Children with ≥1 entry in DCCR indicating an abnormal and disease-causing karyotype | | DCCR |
| Syndromic congenital heart defect | Children with ≥1 entry indicating a recognizable syndrome or a known cause of congenital heart defect<br>ICD-10-codes: Q85, 86, 87, 75.1, 77.1, 79.6, 44.7B, 61.9A | | NPR |
| Maternal use of potentially teratogenic drugs during gestational weeks 3 to 10 | Generic name | ATC codes | DNPR |
| | Antineoplastic and Immunemodulatory agents<br>Warfarin<br>ACE inhibitors<br>Lithium<br>Isotretinoin<br>Misoprostol<br><br>Mifepristone<br>Fluconazole<br>Propylthiuracil<br>Fluoxetine<br>Paroxetine<br>Anti-epileptics | L01-L04<br>B01AA03<br>C09A, C09B<br>N05AN<br>D10BA<br>A02BB01,<br>G02AD06<br>G03XB01<br>J02AC01<br>H03BA02<br>N06AB03<br>N06AB05<br>N03 | |
| Malformation caused by viral infection during pregnancy | Mother diagnosed with viral infection during pregnancy: ICD-10 code O353 or offspring diagnosed with malformation caused by rubella virus: ICD-10 code P350 | | NPR |
| Pre-gestational diabetes | ≥2 filled prescriptions of antidiabetic drug or ≥ 1 ICD code in NPR (ICD 10 codes E10-14, ICD 8 codes 249, 250 or ATC A10 [except A10BA02]) during 6 weeks prior to conception to gestational week 10 | | NPR, DNPR |

| | | |
|---|---|---|
| IVF treatment | Individuals with an entry in the IVF-register with transfer of an embryo within (+/-) 4 weeks of estimated date of conception (calculated based on the MBR) were identified as IVF-users.<br>The specific date of conception in IVF pregnancies was estimated as the embryo transfer date + 3 days if available or 14 days after the beginning of the last menstrual period. | MBR, IVFR |
| Non-cardiac malformation | Non-cardiac birth defects were defined by using ICD-10 codes Q00–Q19 and Q27–Q89 (excluding Q893) and did not include minor birth defects | NPR, CDR |
| Missing data from the first DNBC interview | Mothers who did not answer on the first interview during their pregnancy | DNBC |
| Abbreviations: No.: number, NPR: National Patient Register, MBR: Medical Birth Register, CDR: Danish Register of Causes of Death, DCCR: Danish Cytogenetic Central Register, DNPR: The National Prescription Register, ICD: International Classification of Diseases, ATC: Anatomical Therapeutical Codes, DNBC: Danish National Birth cohort | | |
